# Preterm birth and paternal mental health during the perinatal period: A systematic review protocol

**DOI:** 10.1101/2024.06.13.24308911

**Authors:** Lloyd Frank Philpott, Tim Torsy, Florence D’haenens, Ellen Van denhaute, Inge Tency

## Abstract

**Introduction:** Despite the evidence that fatherhood has a long-term positive and protective effect on men’s health, there is also evidence that fatherhood in the perinatal period can be complex and demanding. The challenging nature of fatherhood during the perinatal period can be further compounded by the experience of preterm birth. Preterm birth, defined as a birth occurring before 37 weeks of gestation, has been identified as a risk factor for maternal stress, anxiety, depression and post-traumatic stress. It can be hypothesised that preterm birth is also a risk factor for fathers mental health; however, as no attempt has been made to systematically review studies that have examined preterm births and father’s mental health during the perinatal period it is currently not known how common or signficant the impact of a preterm birth is on father’s mental health during this life stage.

**Aim:** The aim of the systematic review will be to critically appraise the empirical evidence that examined preterm birth and father’s mental health during the perinatal period.

**Design:** Systematic review

**Methods:** The review will be guided by the PRISMA reporting process. Electronic databases Medline, CINAHL, the Cochrane Library, PsycARTICLES, PsycINFO, Psychology and Behavioural Sciences Collections will be searched to identify studies that meet the inclusion criteria. Studies that researched preterm birth during the perinatal period will be included if fathers mental health was the principal focus of the research, if fathers mental health was in the title and/or aim of the study or if fathers mental health was an outcome or dependent variable. Data will be extracted and presented in narrative form including tables and figures.

**Trial registration:** Trial registration: the protocol for this systematic review has been registered in PROSPERO [**CRD42024536317**].

https://www.crd.york.ac.uk/prospero/display_record.php?RecordID=536317

## Introduction

The perinatal period, which covers the time when a man’s partner becomes pregnant through to the first year after birth is marked by significant change and the absence of routine [1]. While most of these changes are expected and welcome, others can be unanticipated [2]. Despite the evidence that fatherhood has a long-term positive and protective effect on men’s health, there is also evidence that fatherhood in the perinatal period can be complex and demanding [3]. Fatherhood, even when desirable and planned, can be challenging [4]. The challenging nature of fatherhood during the perinatal period can be further compounded by the experience of preterm birth (PTB) [5].

PTB, defined as a birth occurring before 37 completed weeks of gestation, is a major perinatal health problem and the leading cause in perinatal morbidity and mortality worldwide [6,7]. The PTB rate is estimated at 11% or 15 million annually worldwide, [6], with a prevalence varying from 4% to 18% [6].

Preterm birth is one of the most common reason for admission to a neonatal intensive care unit (NICU) and means the start of an emotional, difficult and stressful period for parents [5]. Preterm birth has been associated with symptoms of depression, anxiety and post-traumatic stress [8]. A recent meta-analyse found a higher risk for adverse mental health outcomes for mothers of preterm infants [9]. Preterm birth has also an impact on the mental health of fathers [10]; however, as no attempt has been made to systematically review studies that have examined preterm births and father’s mental health during the perinatal period it is currently not known how common or signficant the impact of a preterm birth is on father’s mental health during this life stage.

To date, research examining paternal perinatal mental health has primarily focused on depression, with less attention given to stress and anxiety [1]. More recently, there has been a shift towards focusing on stress and anxiety, the evidence suggests all three are highly correlated [11]. Other areas of mental health that have received little attention regarding fathers during the perinatal period include Post Traumatic Stress Disorder (PTSD), Obsessive Compulsive Disorder (OCD), and Substance Use [1].

Reported rates of adverse paternal perinatal mental health outcomes are significant; however, they are likely to be under representative of the true prevalence as there is a gendered context in men underreporting mental health symptoms, leading to underdiagnoses [1]. Despite the potential of underreporting the prevalence rates represents a significant public health concern. To date, systematic reviews have been completed on paternal perinatal stress, and anxiety a depression with prevalence rates reported between 6% to 68.8%, 2.4% to 51% and 8.4% respectively. [11, 12, 13]. Risk factors associated with adverse paternal perinatal mental health have also been investigated. Philpott [1] suggests that understanding risk factors helps identity fathers who are likely to develop adverse mental health outcomes during the perinatal period and the evidence suggests that adverse outcomes can relate to personal, maternal, infant, relationship, socioeconomic and environmental factors. One such risk factor is having a preterm birth/infant [10]. To date, no attempt has been made to systematically review studies that have examined preterm births and father’s mental health during the perinatal period; therefore, it is currently not known how common or signficant the impact of a preterm birth is on father’s mental health during this life stage.

A systematic review will give a better understanding of the impact that preterm birth has on father’s mental health during this life stage and will help identify risk populations of fathers, inform service planning and lead to more targeted interventions to support fathers. It will also provide researchers with an opportunity to identify areas where future research is needed. Given the lack of clarity around the current knowledge and the potential impact of preterm birth on father’s mental health, a systematic review is both timely and warranted.

To our knowledge, no systematic review to date has summarized the evidence from studies assessing PTB and adverse paternal mental health outcomes during the perinatal period. Therefore, the aim of this systematic review was to identify, critically evaluate and summarise studies that explored PTB and paternal mental health during the perinatal period.

The objectives for this review will be to identify a) what paternal mental health outcomes have been assessed; b) how paternal mental health outcomes were measured; c) the levels/prevalence of paternal mental health outcomes; d) the impact of paternal mental health outcomes in relation to their health and social relationships and e) interventions and strategies used to manage paternal mental health outcomes. We hypothesize that the association between preterm birth and adverse mental health of fathers during the perinatal period will be confirmed, but methodological differences among the studies might impact on the results.

## Methods

### Study design

A systematic review of peer reviewed literature with respect of paternal mental health during the perinatal period, if possible a meta-analysis, will be performed in accordance with the Preferred Report Items for Systematic Reviews and Meta-analyses (PRISMA) statement. The PRISMA protocols (PRISMA – P) checklist [16] was used to prepare this protocol (S1 table).

### Eligibility criteria and information sources

Inclusion and exclusion criteria are listed in table 1. Electronic searches of literature will be carried out using the following databases: Medline, CINAHL, the Cochrane Library, PsycARTICLES, PsycINFO, Psychology and Behavioural Sciences Collections. No date limit will be applied to the searches.

**Table 1:** inclusion and exclusion criteria Search strategy.

|  | **Inclusion** | **Exclusion** |
| --- | --- | --- |
| **Population** | Fathers in the perinatal period | Fathers not in the perinatal period |
| Design | Quantitative studies of any design | Qualitative studies of any design |
| Language | English | Non-English |
| Availability | Full text available | No full text available |
| Assessment | Peer reviewed articles | Non-peer reviewed articles, grey literature |
| Methods (intervention) | Preterm birth during the perinatal period if: | preterm birth and fathers' mental health was not the |
|  | • fathers' mental health was the principal focus of the research<br>• fathers' mental health was in the title and/or aim of the study<br>• and fathers' mental health was an outcome or dependent variable | main focus of the research<br>• if only women were researched or if the data specific to men and women were not reported separately<br>• if fathers' mental health is not an outcome or a variable<br>• if conducted after or before the perinatal period |

### Search strategy

Search terms to be used are listed in table 2. The search strategy will include the Boolean terms “OR”/ “AND” and truncation (*). Phrase searching (““) will also be used. Keywords and search terms are demonstrated in Table 2.

**Table 2:** list of search terms to be applied.

| Intervention terms | Search terms |
| --- | --- |
| Population: Father | father* OR paternal OR dad* OR male OR men OR partner |
| Population: Preterm | 'preterm infant' OR 'premature infant' OR 'preterm neonate' OR 'prematurity' OR 'low birth weight infant' OR 'very low birth weight infant' OR 'preterm birth' OR 'premature' OR 'preterm labor' OR 'gestational age' OR 'low birth weight' OR 'neonatal intensive care unit' OR NICU OR neonatology OR 'neonatal unit' |
| Context: Perinatal | prenatal OR prepartum OR antenatal OR antepartum OR perinatal OR peripartum OR postnatal OR postpartum OR preg* OR childbirth OR birth OR labour OR labor |
| Outcome: Mental health | 'mental health' OR 'mental disorders' OR 'mental health services' OR 'adjustment disorders' OR 'mental illness' OR 'mental disease' OR 'mental problem' OR 'adverse mental health' OR 'mental health symptoms' OR distress OR psychiat* OR 'mental health wellbeing' OR 'mental health well-being' OR 'stress' OR 'Anxiety' OR 'Depression' OR 'Post Traumatic Stress Disorder' OR 'PTSD' OR 'Obsessive Compulsive Disorder' OR 'OCD' OR 'Substance abuse' |

**Table 3:**
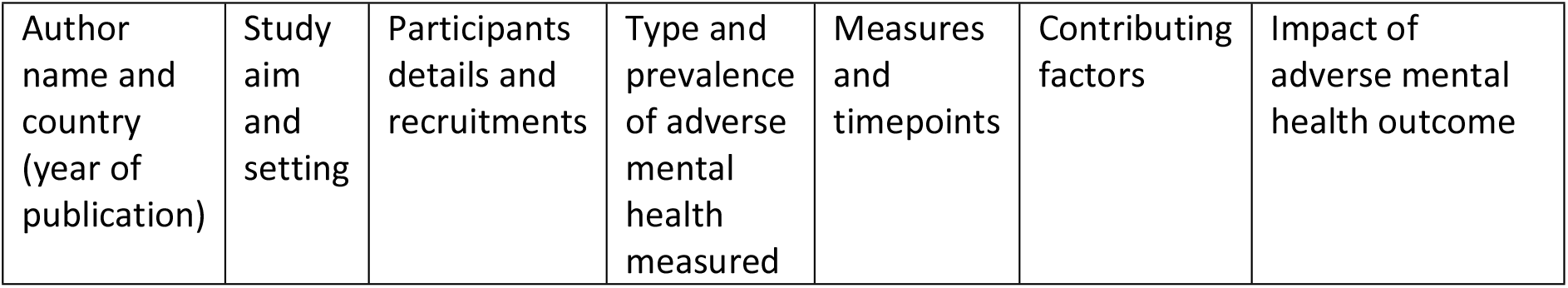
Characteristics of each study.

**Table 4:**
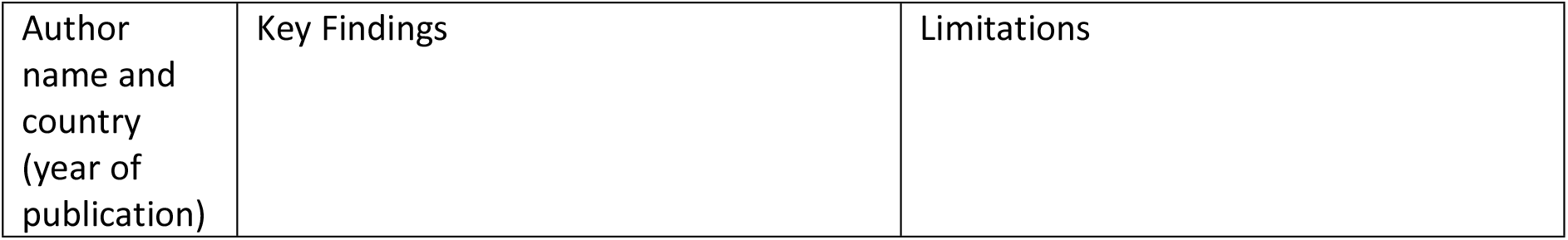
Findings and limitations of each study Data synthesis.

### Study selection

Relevant studies will be identified through electronic database searching, which utilizes the search terms. The relevant studies from the databases will be exported to Covidence and the duplicates will be removed. Two independent researchers (LP and IT) will initially screen the title and abstracts of the studies. The studies will be removed if they do not fit the inclusion criteria. After that, the full text of the remaining studies will be reviewed by two independent researchers (LP and IT) to choose the studies for the review. If there are any discrepancies, a third researcher (TT) will assess the studies to make the final decision. The study selection will be regulated and exhibited using a PRISMA flow diagram (Fig 1).

### Quality appraisal and risk of bias assessment

The methodological quality of the included studies will be assessed by two researchers (EvD and FD’n) using the Joanna Briggs Institute (JBI) critical appraisal checklist for systematic review and research synthesis [15] and reported as per the Preferred Reporting Items for Systematic Reviews and Meta-analyses (PRISMA) [14]. Discrepancies will be resolved through mutual discussion and agreement. Considering the type of studies that will be included, various tools for assessing the risk of bias will be utilised.

### Data collection

One author (LP) will extract the relevant data from the studies: author name and country, year of publication, study aim, study setting, study design, participant details and recruitments, type of adverse mental health outcome, findings and limitations. Another author will independently cross-check data extraction (IT). Discrepancies will be resolved by consensus. The data extraction tables are presented in Tables 1 (characteristics of each study) and Table 2 (findings and limitations of each study). Data analysis will be descriptive. If the nature of the included studies allows, a meta-analysis will be undertaken. If the criteria for a meta-analysis are not met (i.e., heterogeneity in research designs, interventions, and outcome measures among studies) a narrative synthesis of the findings will be undertaken.

The abstracted data from the selected studies will be synthesized and presented according to the primary aim and objectives of the review. Extracted data from eligible studies will be presented in evidence tables and a narrative synthesis will provide a summary of the prevalence of adverse paternal mental health related to preterm birth in the perinatal period, the contributing factors to adverse mental health outcomes, the measurement tools used to assess adverse mental health outcomes, the time point/s when adverse mental health outcomes were measured during the perinatal period and what interventions/strategies are available for fathers to manage and address adverse mental health outcomes.

### Ethics and dissemination

The review will not require ethical approval and the findings will be published in a peer reviewed journal using the PRISMA guidelines and presented orally at an International research conference.

## Discussion

Fatherhood can be complex and challenging [4], in particular when experiencing a PTB [5]. PTB has been associated with symptoms of depression, anxiety and stress [8] However, systematic reviews about the impact of PTB on father’s mental health are lacking.

Therefore, this review aimed to identify, critically evaluate and summarize studies that explored PTB and paternal mental health and wellbeing during the perinatal period, including outcomes, measuring instruments, prevalence, impact and interventions. The rigorous and transparent methodology of this systematic review will result in a better understanding of the potential impact of PTB on fathers’ mental health and lead to more targeted interventions to support fathers during this life stage.

## Data Availability

No datasets were generated or analysed during the current study. All relevant data from this study will be made available upon study completion

## Abbreviations

JBI: Joanna Briggs Institute
NICU: Neonatal Intensive Care Unit
OCD: Obsessive Compulsive Disorder
PTB: Preterm Birth
PTSS: Post Traumatic Stress Disorder

## Supporting information

S1 Table. PRISMA-P 2015 checklist.

S2 JBI CRITICAL APPRAISAL CHECKLIST FOR SYSTEMATIC REVIEWS AND RESEARCH SYNTHESES

## Author contributions

**Conceptualization:** LP and IT

**Investigation:** LP, IT, TT, EvD and FD’h

**Methodology:** LP, IT, TT, EvD and FD’h

**Project administration:** LP anPLOS one d IT

**Supervision:** LP and IT

**Validation:** LP, IT, TT, EvD and FD’h

**Writing – original draft:** LP, IT, TT, EvD and FD’h

**Writing – review & editing:** LP, IT, TT, EvD and FD’h

